# Predictive models for starting antiseizure medication withdrawal following epilepsy surgery in adults

**DOI:** 10.1101/2022.07.22.22277802

**Authors:** Carolina Ferreira-Atuesta, Jane de Tisi, Andrew W. McEvoy, Anna Miserocchi, Jean Khoury, Ruta Yardi, Deborah T. Vegh, James Butler, Hamin J. Lee, Victoria Deli-Peri, Yi Yao, Feng-Peng Wang, Xiao-Bin Zhang, Lubna Shakhatreh, Pakeeran Siriratnam, Andrew Neal, Arjune Sen, Maggie Tristram, Elizabeth Varghese, Wendy Biney, William P Gray, Ana Rita Peralta, Alexandre Rainha-Campos, António JC. Gonçalves-Ferreira, José Pimentel, Juan Fernando Arias, Samuel Terman M, Robert Terziev, Herm J. Lamberink, Kees P.J. Braun, Willem M Otte, Fergus J. Rugg-Gunn, Walter Gonzalez, Carla Bentes, Khalid Hamandi, Terence J. O’ Brien, Piero Perucca, Chen Yao, Richard J. Burman, Lara Jehi, John S. Duncan, Josemir W Sander, Matthias Koepp, Marian Galovic

## Abstract

More than half of adults with epilepsy undergoing resective epilepsy surgery achieve long-term seizure freedom and might consider withdrawing antiseizure medications (ASMs). We aimed to identify predictors of seizure recurrence after starting postoperative ASM withdrawal and develop and validate predictive models.

We performed an international multicentre observational cohort study in nine tertiary epilepsy referral centres. We included 850 adults who started ASM withdrawal following resective epilepsy surgery and were free of seizures other than focal non-motor aware seizures (auras) before starting ASM withdrawal. We developed a model predicting recurrent seizures, other than auras, using Cox proportional hazards regression in a derivation cohort (n=231). Independent predictors of seizure recurrence, other than auras, following the start of ASM withdrawal were focal-aware seizures after surgery and before withdrawal (adjusted hazards ratio [aHR] 5.5, 95% confidence interval [CI] 2.7-11.1), history of focal to bilateral tonic-clonic seizures before surgery (aHR 1.6, 95% CI 0.9-2.8), time from surgery to the start of ASM withdrawal (aHR 0.9, 95% CI 0.8-0.9), and number of ASMs at time of surgery (aHR 1.2, 95% CI 0.9-1.6). Model discrimination showed a concordance statistic of 0.67 (95% CI 0.63-0.71) in the external validation cohorts (n=500). A secondary model predicting recurrence of any seizures (including auras) was developed and validated in a subgroup that did not have auras before withdrawal (n=639), showing a concordance statistic of 0.68 (95% CI 0.64-0.72). Calibration plots indicated high agreement of predicted and observed outcomes for both models.

We show that simple algorithms, available as graphical nomograms and online tools (predictepilepsy.github.io), can provide probabilities of seizure outcomes after starting postoperative ASMs withdrawal. These multicentre-validated models may assist clinicians when discussing ASM withdrawal after surgery with their patients.

## Introduction

More than half of adults with drug-resistant epilepsy who undergo resective surgery achieve postoperative seizure freedom.^1^ After successful surgery, they and their treating clinicians need to decide whether antiseizure medication (ASM) should be reduced or withdrawn. Continued administration of ASMs may have side effects, teratogenic implications and increase healthcare costs.^2^ Conversely, ASM withdrawal might increase the risk of seizure relapse and subsequent injuries, epilepsy-related mortality, including SUDEP, occupational and driving constraints, and social stigma.^3,4^ Several studies have identified risk factors for seizure recurrence following ASM withdrawal after epilepsy surgery. These include, among others, epilepsy duration at the time of surgery, characteristics of presurgical seizures, preoperative MRI abnormalities, the timing of ASM withdrawal, incomplete resection, and postoperative EEG abnormalities.^5–12^

Recent studies have shown the feasibility of predicting outcomes of ASM withdrawal in non-surgical cases,^13^ and after paediatric epilepsy surgery.^11^ A prognostic model synthesizing the clinical characteristics predicting recurrence after ASM withdrawal following successful epilepsy surgery in adults is not yet available.^14^ The lack of tools to guide ASM withdrawal decisions leads to significant heterogeneity in the timing and strategies of drug discontinuation.^8,9,12,13,15,16^ There is a need for practical instruments to support these clinical decisions and inform individuals and their relatives about realistic expectations of the risks and outcomes following ASM withdrawal.

We aimed to develop and validate a prediction model that provides probabilities of seizure recurrence following ASM withdrawal after epilepsy surgery in adults with readily available clinical variables.

## Methods

We developed the prediction model using prospectively collected baseline and follow-up data from an ongoing consecutive registry of individuals who had epilepsy surgery at a tertiary centre in London (United Kingdom). The cohort has been reported previously in detail.^1^ All participants were prospectively followed in yearly intervals (median total follow up duration 11 years, interquartile range [IQR] 6 – 16 years). Annual postsurgical seizure outcome was determined using the International League Against Epilepsy (ILAE) outcome scale,^17^ and the participants’ medication regime was noted. Data on ASM reductions’ timing was corroborated by reviewing medical notes and extracting the exact timing of starting ASM withdrawal and seizure recurrences.

We randomly divided the London cohort into derivation and internal validation cohorts with a 2:1 ratio for model development and internal validation. For the development of the model, we only used the derivation cohort.^18^

For external model validation, we collected baseline and follow-up data of individuals undergoing epilepsy surgery with at least one year of postsurgical follow up at eight tertiary epilepsy referral centres (Cleveland [United States], Cape Town [South Africa], Shenzhen [China], Oxford [United Kingdom], Melbourne [Australia], Cardiff [United Kingdom], Lisbon [Portugal], and Bogotá [Colombia]). A detailed description of each cohort and the informed consent procedures are described in the Online Supplement.

In all cohorts, we included consecutive adults who underwent resective epilepsy surgery, had at least one year of postoperative follow-up, had no seizures other than focal non-motor aware seizures after surgery, i.e. ILAE outcome class 1 or 2,^17^ and started reducing ASMs. We excluded those who did not achieve initial seizure freedom other than focal-aware seizures (ILAE outcome class 3 or worse), did not attempt ASM withdrawal, had disconnective procedures, multiple brain surgeries, or had insufficient follow-up data. Acute postoperative seizures (i.e., occurring during the first 30 days after surgery) were not considered to be seizure recurrences.^19^ Data for model development was complete.

### Outcomes

The primary model outcome was time to seizure recurrence other than focal non-motor aware seizures (auras, i.e., ILAE outcome Class 3-6)^17^ after the start of ASM withdrawal. Due to several considerations, we only considered seizures other than focal non-motor aware seizures, i.e. seizures with motor symptoms or those associated with impaired awareness. Firstly, the presence of focal non-motor aware seizures only may not be regarded as a poor outcome.^20^ Seizures other than focal non-motor aware seizures are arguably clinically more relevant than focal non-motor aware seizures because they are more likely to cause injuries and lead to increased morbidity and mortality.^21^ Secondly, individuals with only focal non-motor aware seizures may still consider ASM withdrawal and thus, including them makes the model more applicable.^22^ Third, in some jurisdictions, focal non-motor aware seizures do not preclude driving.

On the other hand, given that focal non-motor aware seizures may have an impact on quality of life and represent a red flag for starting ASM discontinuation,^23^ we developed a secondary model to predict any type of seizure recurrence (i.e., ILAE outcome Class 2-6) that included individuals that were completely seizure-free after surgery and before withdrawal (i.e. did not have any focal non-motor aware seizures).

As an additional outcome, we also assessed the time to complete withdrawal of all ASMs.

### Development of the primary model

We performed a literature review of previously-reported predictors of seizure recurrence after ASM withdrawal following epilepsy surgery (see online Supplement). We chose predictors consistently reported to have a significant and independent association with the outcome and easily ascertained in different settings with varying clinical expertise. The selected predictors are also part of the routine diagnostic tests for people who ultimately undergo epilepsy surgery. We did not include data on postsurgical electroencephalography (EEG) because there were insufficient available data,^24^ as this was not routinely performed in several centres involved in this study.^25^ Data on percentage and rate of dose reduction, and reasons to halt withdrawal other than seizure relapse were not available.

Using Kaplan-Meier plots, we estimated the proportion of individuals remaining seizure-free at various time-points after ASM withdrawal commencement. We used univariable Cox proportional hazards regression analyses to assess the relevance of previously reported variables and identify any other potential predictors. Hazard ratios (HRs) were estimated with 95% confidence intervals (CI). The model was censored at the recurrence of a seizure other than focal non-motor aware seizures or on the last follow-up day.

Variables previously reported were included in the multivariable analyses and any significant variable (p<0.05) in the univariable analyses. The multivariable model was simplified by backward stepwise elimination based on the Akaike Information Criterion (AIC).^26^ The AIC evaluates the fit of a model while penalizing overfitting and provides a means to select the most relevant variables regardless of their p-value. Lower AIC indicates a better fit, i.e., a higher likelihood with fewer parameters.

We checked the statistical assumptions for Cox proportional hazard regressions and they were fulfilled.

### Validation of the model

The performance of the model in the internal and external validation cohorts was assessed using discrimination and calibration.^27^ Discrimination refers to how well the model distinguishes between participants with favorable or unfavorable outcomes. We used the concordance (c) statistic to measure discrimination, which corresponds to the area under the receiver operating characteristic (ROC) curve. Calibration indicates the agreement between outcomes that were predicted by the model and those that were observed. We used calibration curves, which plot the predicted risk given by the model against the observed risk, to assess calibration.

Prognostic models obtained from multivariable regression analyses can be overfitted and overestimate predictions when applied to a new cohort.^28^ To get realistic precision estimates and confidence intervals, bootstrapping techniques were applied. A shrinkage factor was estimated from 1000 random samples to correct the c statistic for over-optimism, and 95% confidence intervals for risk estimates were generated to account for residual uncertainty. Internal-external cross-validation was performed to evaluate the model across different populations, as described previously.^18^ The final AIC value was calculated over the pooled data set.^18^

### Model predictions

The final risk estimates were estimated using combined data from all cohorts to increase generalisability.^18^ To improve the practical usability of the model, we generated an easily estimated nomogram, a two-dimensional diagram that allows the graphical computation of a mathematical function. We also developed an interactive, user-friendly, and convenient web tool that provides individualized outcome estimates with corresponding 95% CIs and graphical representation.^29^

### Secondary models

A secondary model was developed, including only those completely seizure-free after surgery (ILAE Class 1), i.e. those that did not have any postsurgical focal non-motor aware seizures. The secondary outcome parameter for these analyses was complete seizure-freedom, i.e. counting focal non-motor aware seizures as seizure relapses. As a sensitivity analysis, we also created a similar model that only included those undergoing temporal lobe surgery. We also developed a model of time to withdrawal of all ASMs in individuals that were completely seizure free after surgery. The same methodologies as described above were applied, and data was assessed and cross-validated in the combined cohort.

Development and validation of the presented models followed established recommendations (i.e. TRIPOD).^30^ Two-sided p-values < 0.05 were considered statistically significant. Analyses were performed and updated using R version 3.6.2 using the packages “survival”, “survminer,” “rms”, “survivalROC”.

### Data availability

The data that support the findings of this study are available from the corresponding author, upon reasonable request.

## Results

### Participant characteristics

The London cohort included 350 adults, of whom 231 randomly selected were used for model development (derivation cohort) and 119 for internal validation (internal validation cohort). External cohorts included 500 adults (Cleveland [*n*=98], Cape Town [n=105], Shenzhen [*n*=83], Melbourne [*n*=48], Cardiff [*n*=44], Lisbon [*n*=42], Oxford *[n*=40], Bogota [*n*=40]). We included 850 subjects overall, all of whom were seizure-free other than focal non-motor aware seizures between surgery and the start of ASM withdrawal (ILAE outcome 1 or 2, Supplementary Figure 1).

The overall clinical and demographic characteristics are provided in Supplementary Table 1. In the combined data set, the median time between surgery and the start of ASM withdrawal was 1.0 years (*IQR* 0.5-2.2). Kaplan Meier estimates indicate that 80% remained free from seizures other than focal non-motor aware seizures two years after starting ASM withdrawal and 72% after four years (Figure 1). At the end of follow-up, 317 (37%) participants had experienced a seizure relapse (including focal non-motor aware seizures), of whom 47 only had focal non-motor aware seizures. 308 (36%) individuals ultimately came off all ASMs. The median time between the start of ASM withdrawal to the complete withdrawal of all ASMs was 1.5 years (*IQR* 0.44-2.83).

**Figure 1.**
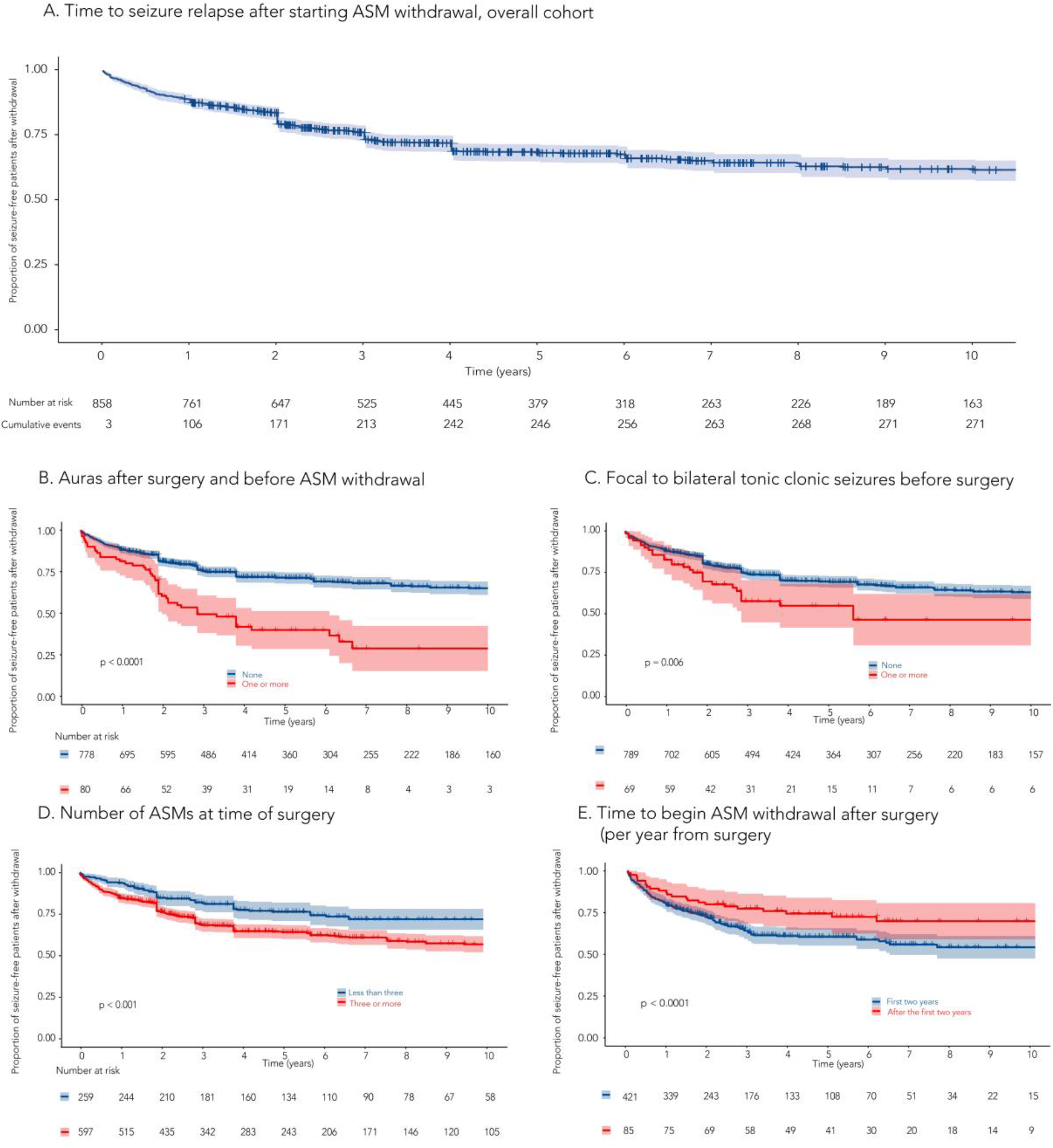
Impact of predictors on Kaplan-Meier estimates (time to non-aura seizure recurrence after starting ASM withdrawal) Panel **A** represents the Kaplan-Meier estimates of time to non-aura seizure recurrence in the overall cohort (n=850). Panels **B-E** show the impact of predictors included in the final model on the time to non-aura seizure recurrence. Time at baseline was beginning of ASM withdrawal. Shaded band represents 95% confidence interval.

### Primary model

Variables consistently described as relapse predictors (time between surgery and beginning of ASM withdrawal, history of FBTCS before surgery, number of ASMs at time of surgery, duration of epilepsy at time of surgery, hippocampal sclerosis on neuropathology, incomplete resection of an epileptogenic lesion, extratemporal lobe surgery, normal MRI before surgery, and presurgical seizure frequency) were included in the multivariable regression. The univariable analysis showed that postsurgical focal non-motor aware seizures before starting withdrawal were significantly associated with relapse after starting ASM withdrawal (Supplementary Table 2), and thus were also included.

After simplification based on the AIC, four predictors remained in the final multivariable model (Table 1A): focal non-motor aware seizures after surgery and before the starting ASM withdrawal, history of FBTCS, the time between surgery and starting withdrawal, and ASMs number at the time of surgery. Figure 1 displays the impact of these predictors on time to non-aura seizure relapse after starting ASM withdrawal in the combined data of all cohorts.

**Table 1.** Multivariable Cox regression analysis of time to non-aura recurrence reccurence after beginning of ASM withdrawal following epilepsy surgery.

| Predictors | aHR (95% CI) | p-value | $\Delta$<br>AIC* |
| --- | --- | --- | --- |
| Auras after surgery before beginning ASM withdrawal | 5.53 (2.74-11.15) | <0.0001 | -14.1 |
| Time to beginning of ASM withdrawal (*per year from surgery*) | 0.90 (0.82-0.98) | 0.02 | -4.8 |
| Focal to bilateral tonic- clonic seizures before surgery | 1.60 (0.91-2.82) | 0.09 | -1.0 |
| Number of ASMs at time of surgery | 1.24 (0.95-1.60) | 0.10 | -0.5 |
| Normal presurgical MRI | eliminated step 5 | 0.45 | 1.5 |
| Age at surgery | eliminated step 4 | 0.59 | 1.7 |
| Presurgical seizure frequency (*as an ordinal scale*) | eliminated step 3 | 0.60 | 1.7 |
| Hippocampal sclerosis on neuropathology | eliminated step 2 | 0.67 | 1.8 |
| Duration of epilepsy at time of surgery | eliminated step 1 | 0.74 | 1.9 |

N=231. aHR, adjusted hazard ratio; MRI, magnetic resonance imaging; $\Delta$ AIC=change in Akaike information criterion after elimination of a variable at each step; \* A negative value implies that the variable improves the model and should be kept in the model.

The model had an optimism-corrected *c* statistic of 0.68 (*95% CI* 0.58-0.79) in the internal validation (*n*=119) and 0.67 (*95% CI* 0.63-0.71) in the external validation (*n*=500) cohorts. Calibration plots indicated high agreement between predicted and observed data in the internal and external validation cohorts (Supplementary Figure 2). Internal-external cross-validation showed that the c statistic remained stable across different populations (Supplementary Table 3).

The model, named *WAMS* (<u>W</u>ithdrawal of <u>A</u>ntiseizure <u>M</u>edication After <u>S</u>urgery) was translated into an easy-to-use graphical nomogram (Figure 3A) predicting outcomes two and four years after starting withdrawal. Time-variable prediction estimates can be estimated using a freely available practical online tool with a graphical user interface on https://predictepilepsy.github.io/

### Secondary models

We performed a secondary analysis looking at predictors of complete seizure-freedom (binary outcome) in postoperatively seizure-free participants who did not have any focal non-motor aware seizures (ILAE Class 1, *n*=639). The resulting model (Figure 2, Supplementary Tables 4, 5) included the following predictors: history of FBTCS before surgery, presurgical seizure frequency, the time between surgery and starting withdrawal, duration of epilepsy before surgery and history of febrile seizure. The model showed an optimism-corrected *c* statistic of 0.68 (*95% CI* 0.64-0.72) and a high agreement between predicted and observed data (Supplementary Figure 2). Prediction estimates can be determined using a graphical nomogram (Figure 3B) or the online tool. The results for a similar model in people undergoing temporal lobe surgery can be found in the online supplement (Supplementary Tables 6, 7; Supplementary Figures 2, 3).

**Figure 2.**
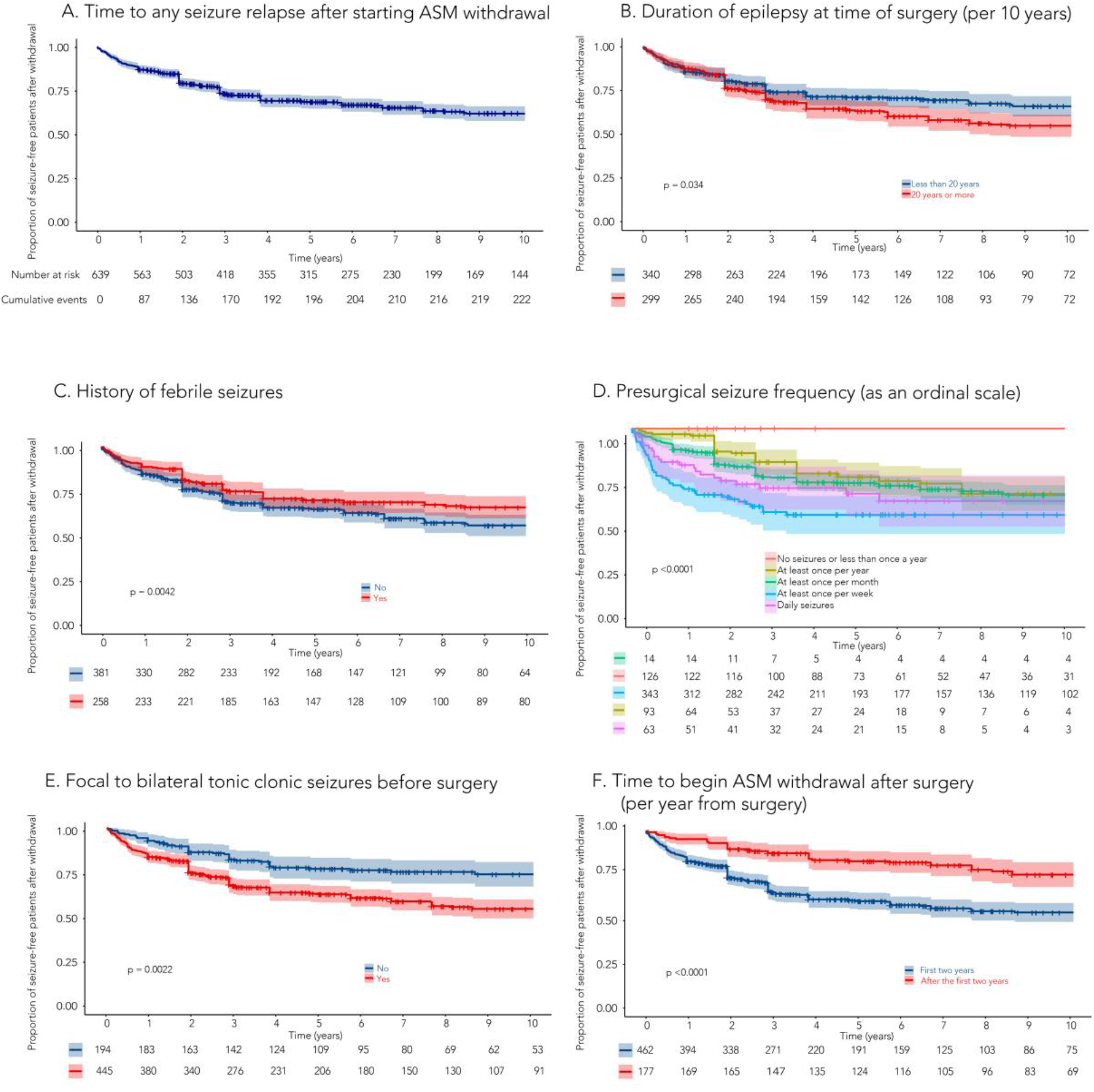
Impact of predictors on Kaplan-Meier estimates (time to any seizure recurrence after starting ASM withdrawal) Panel A represents the Kaplan-Meier estimates of time to any seizure recurrence in the completely seizure-free cohort (n=639). Panels **B-F** show the impact of predictors included in the final model on the time to any seizure recurrence. Time at baseline was beginning of ASM withdrawal. Shaded band represents 95% confidence interval.

**Figure 3.**
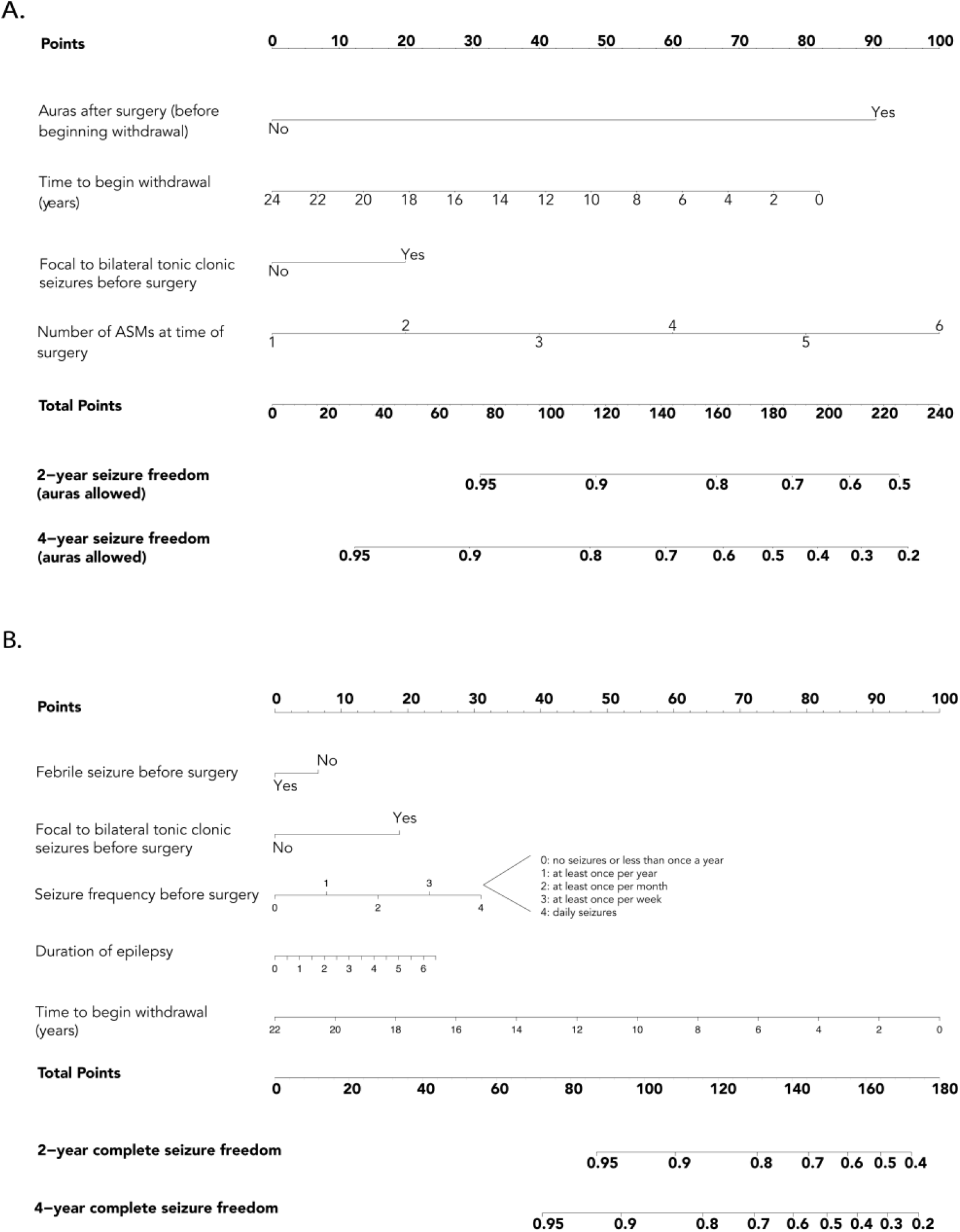
Nomograms for predicting 2- and 4-year seizure outcome after starting ASM withdrawal following epilepsy surgery. Freedom from non-aura seizures (**A**) and from any seizures including focal non-motor aware seizures (**B**) after starting ASM withdrawal after epilepsy surgery. Instructions: Determine the individual risk in three steps: 1) For every variable on the left, count the points given at the top, 2) Add up the points to a total, 3) Determine the associated recurrence risk at 2 and 4 years according to the calculated point total.

The results of a model predicting the withdrawal of all ASMs are displayed in the online supplement (Supplementary Tables 8, 9; Supplementary Figure 2). This model had an optimism-corrected *c* statistic of 0.73 (*95% CI* 0.68-0.78) and can be calculated using a graphical nomogram (Supplementary Figure 4).

ROC curves for all models are displayed in Supplementary Figure 5.

### Case simulation

Examples of how to use the nomograms and online tools based on two fictional cases are illustrated in Figure 4.

**Figure 4.**
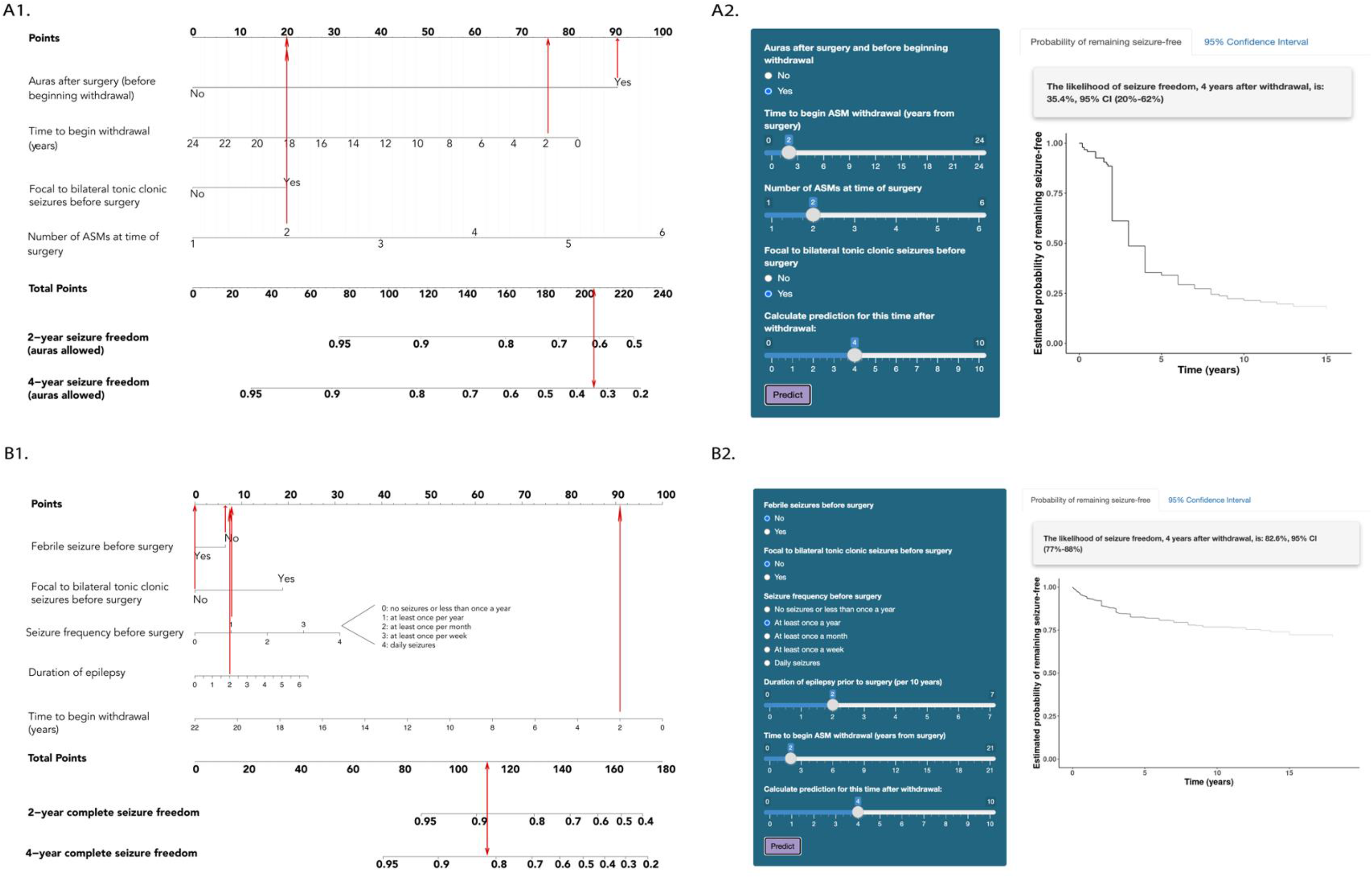
Fictional case simulations. Examples on how to use the nomogram (**A1, B1**) and online tool (**A2, B2**) based on two fictional cases reflecting real life scenarios. Case A (panels **A1-2**) is a 35-year-old individual with a 20-year history of epilepsy with 1-2 preoperative FBTCS per year, no febrile seizures, taking two ASMs at time of surgery, and focal non-motor aware seizures after surgery, who is considering withdrawal of ASMs two years after epilepsy surgery. The models show a low chance of remaining free from non-aura seizures four years after starting withdrawal (35%, 95% CI 20-62). Case B (panels B1-2) is an individual with similar characteristics but no history of FBTCS and complete postoperative seizure-freedom (i.e. no focal non-motor aware seizures after surgery). The models predict a higher chance of remaining seizure-free (freedom from non-aura seizures after four years 86%, 95% CI 80-92; complete seizure freedom after four years 83%, 95% CI 77-88).

## Discussion

The decision to withdraw, or even just to reduce, ASMs after having reached seizure-freedom following epilepsy surgery is a common clinical problem. Here, we developed and validated predictive models that provide individualized probabilities of seizure outcomes in people that have started ASM withdrawal after epilepsy surgery. The models will provide objective recurrence expectations for people with epilepsy considering ASM withdrawal. They could assist presurgical counselling and decision making, especially in individuals who prioritize ASM withdrawal as a marker of surgical success. The models can be easily estimated using graphical nomograms or a freely available online tool.

The included predictors are clinically meaningful. We identified the timing of ASM withdrawal after surgery as a significant predictor in all models. This is relevant because the optimal time for starting ASM withdrawal after successful surgery is unknown, and approaches are heterogeneous, mainly relying on the clinician’s personal experience and subjective risk assessment.^31–34^ In the context of non-surgical individuals, a common practice is to consider ASM reduction after two years of seizure-freedom. Despite robust evidence, this approach is often implemented in postsurgical cases.^3,14,15,33^ Some studies found that early withdrawal (<2 years) is associated with an increased risk of relapse compared with later withdrawal.^8,9,13,16,34^ Others reported that early withdrawal did not affect long-term outcomes.^35,36^. Our models might support individualized decisions on the timing of ASM withdrawal as we incorporated timing as an adjustable variable. The models will allow clinicians to adjust dynamically and individually postoperative observation time before attempting medication withdrawal according to individual characteristics and preferences. It might be feasible and safe to start withdrawal earlier in low-risk cases and prolong the observation period in high-risk individuals, although such an approach will require further prospective studies.

We found that focal non-motor aware seizures after surgery were the strongest predictor of seizure recurrence following ASM withdrawal. They may represent an early marker of surgical failure due to the incomplete removal of the epileptogenic zone.^1,37^ It has been previously described that entirely seizure-free individuals had a lower risk of seizure relapse with impaired awareness in the following year than individuals with only focal non-motor aware seizures.^37^

Taking more ASMs at the time of surgery, having a higher preoperative seizure frequency, and preoperative FBTCS have been shown to be associated with epilepsy severity and increased risk of seizure relapse in surgical and non-surgical cases.^10,11,37^ FBTCS involve and spread through distributed brain networks and maybe a biomarker of a more diffuse epileptogenic zone that is more difficult to completely resect.^38^ Long presurgical duration of epilepsy^39^and having no history of febrile seizures^40^ have previously been reported as risk factors for poor surgical outcome. These factors could account for a higher risk of seizure relapse when there is an attempt at ASM withdrawal. We found that acute post-surgical seizures, the resection location (temporal versus extratemporal), and the pathological findings were not associated with seizure freedom after starting postsurgical ASM withdrawal. These factors have, however, been previously related to surgical success.^41,42^

This study has several strengths. We evaluated data from one of the largest multicentre populations of people who attempted ASM withdrawal after successful surgery. When validated across nine cohorts from high and low-income countries, the models showed robust performance, supporting their generalizability.^27^ Our models were well calibrated and may, thus, provide realistic statistical estimates of the risk of seizure relapse in individuals that have decided to attempt ASM withdrawal, which is relevant for decision making and follow-up recommendations.^27,29,43^

Data were acquired in the clinical setting of tertiary epilepsy referral centres. Thus, the findings reflect real-life scenarios, and the results can be readily applied to clinical situations. The models were developed using consecutive long-term single-center data from 1990-2016, but the external validation was done in several cohorts with recent data, contributing to temporal validation.^27^ This confirmed the performance of the model in more contemporary settings given changes to the surgical candidates and procedures over the last decades. We conducted a review to identify predictors of outcomes comprehensively evaluated in previous reports. The selected predictors were well defined, easily measured, and routinely available. Additional models predicting complete seizure freedom and the likelihood of achieving complete ASM withdrawal were also developed. We implemented the main models in an online tool that will increase their accessibility and practicality.

Our study was devised in an intention-to-treat manner. We included all participants that decided to start ASM withdrawal, regardless of whether the withdrawal was completed or not. Thus, the models provide the probabilities of seizure recurrence to individuals that are considering to start ASM withdrawal, rather than to those who already successfully reduced doses and are coming off all medications. The participants included in our study had a low pre-test likelihood of recurrence based on clinical expertise.

Developing a predictive model involves making compromises. We did not include predictors not routinely assessed in clinical practice or those that did not support sufficient validation data. Future studies might refine predictions by including data from postsurgical EEG, blood biomarkers, advanced neuroimaging, and genetic data. Such additional biomarkers could further improve model discrimination.

Our study has several limitations. Our results are only applicable to adults, and different models should be used for children.^11^ The models should only be applied when data for each included predictor is available. Due to the cohorts’ observational character, we could not implement a systematic withdrawal procedure. The decision to start withdrawing ASMs was dependent on the participants’ characteristics and preferences, which could lead to increased data variability. This approach reflects a real-life clinical setting and makes the models applicable to various realistic withdrawal protocols. The models should only be applied to those who are already considered potential candidates for ASM withdrawal by their treating physicians.

There were baseline differences between the included cohorts, but this reflects real-life scenarios and supports the model’s generalizability to a wide range of cohorts and settings. Data on missed medications was unavailable in some cohorts, which could account for provoked seizure relapse. Lastly, due to the large variability of treatment regimens, we did not differentiate or adjusted for specific ASMs or percentage and rate of dose reduction. Future controlled studies are needed to identify the impact of different dose reduction protocols with recurrence risks.

In summary, we developed and validated simple algorithms that can help assert decisions on postsurgical ASM withdrawal. They might support individuals and attending physicians by providing quantitative risk estimates of seizure relapse that are dependent on the timing of starting ASM withdrawal and are a step towards more personalized epilepsy care.

## Data Availability

The data that support the findings of this study are available from the corresponding author upon reasonable request.

## Abbreviations

ASM: AntiSeizure Medication
IQR: Interquartile Range
ILAE: the International League Against Epilepsy
HR: Hazard Ratio
aHR: Adjusted Hazard Ratio
MRI: Magnetic Resonance Imaging
EEG: Electroencephalogram
AIC: Akaike Information Criterion
FBTCS: Focal to Bilateral Tonic Clonic Seizures
WAMS: Withdrawal of Antiseizure Medication After Surgery

## Acknowledgements

CFA and MG had full access to all the data in the study and takes responsibility for the integrity of the data and the accuracy of the data analysis. This work was carried out at University College London Hospitals Comprehensive Biomedical Research Centre, which receives a proportion of funding from the UK Department of Health’s National Institute for Health Research centres funding scheme. AS is supported by the NIHR Oxford Biomedical Research Centre. The RACP Foundation Margorie Hooper Scholarship supports AK. JWS receives support from the Dr Marvin Weil Epilepsy Research Fund, UK Epilepsy Society, Christelijke Vereniging voor de Verplegingvan Lijders aan Epilepsie (Netherlands). Special thanks to Harry Marr BSc (Director of Software Engineering at GitHub) for his technical assistance in creating the website. No compensation was provided, and permission granted to include his name.

