## Supplement for "Predictive models for starting antiseizure medication withdrawal following epilepsy surgery in adults"

### Online Supplement

#### Literature review

PubMed, Cochrane, and Ovid were searched and updated periodically until June 2021 to select the articles for review. We identified consistent evidence for eleven predictors of seizure recurrence following postoperative ASM withdrawal: age (at onset and at surgery), epilepsy duration, pre-surgical seizure frequency, history of focal to bilateral tonic-clonic seizures (FBTCS), number of ASMs at surgery, abnormalities on preoperative MRI, location of surgery, incomplete resection of a lesion, pathology findings, postsurgical auras before the onset of ASM withdrawal, and time from surgery to the start of ASM withdrawal.[2–4]
The following keywords were used in all the databases: *epilepsy surgery, antiseizure medication, antiepileptic drug, withdrawal, reduction, seizure outcome, seizure relapse, predictors of relapse, prognosis.* We also searched reference lists of relevant articles, reviews, and meta-analyses for additional sources. We only included original full-text articles in English, Spanish or French. After eliminating duplicates, titles and abstracts, all articles were screened and non-relevant articles were excluded (e.g., case reports; pharmacological, neurosurgical or molecular studies; animal studies). A full-text assessment was carried out, and eligibility was based on the quality and relevance of the evidence.

#### Informed consent procedures

| Cohort name | Ethics Committee /IRB | Decision |
|---|---|---|
| London | Joint Ethics Committee of the National Hospital for Neurology and Neurosurgery and University College London Institute of Neurology | Waived |
| Lisbon | Hospital de Santa Maria Comite de Etica | Waived |
| Bogota | Uniepilepsias, Instituto Roosevelt Comite de Etica | Waived |
| Oxford | Local Clinial Audit, Oxford University Hospital Trust internal committee | Approved |
| Cape Town | University of Cape Town Human Research Ethics Committee | Approved |
| Melbourne | Melbourne Health and Royal Children's Hospital Human Research Ethics Committees | Approved |
| Cardiff | Continuous Service Improvement Office and Vale University Health Board | Approved |
| Shenzhen | Shenzhen Children Hospital Ethics Committee | Waived |

| | | |
|---|---|---|
| Cleveland | Cleveland Clinic's Institutional Review Board | Approved |

#### Description of cohorts

For the London, United Kingdom, cohort, baseline and follow-up data of 968 individuals was collected from a large ongoing prospective longitudinal study[1]. Adults who underwent resective epilepsy surgery between January 1, 1990 and April 1, 2016, had at least one year of postoperative follow-up, were postoperatively free from non-aura seizures, i.e. ILAE outcome class 1 or 2 , and that started antiepileptic drug reduction were included. Those who (i) did not achieve freedom from non-aura seizures before ASM withdrawal (n=512), (ii) did not attempt withdrawal (n=83), (iii) had multiple brain surgeries (n=6), or (iv) had insufficient follow-up data (n=17) were excluded. All participants were postoperatively followed in yearly intervals (median total follow up duration 11 years, interquartile range [IQR] 6 – 16) by a neurology consultant and a clinical data manager. In case they were not reached, their general practitioners were contacted and, in cases of uncertainty, the participants' next of kin, as described previously.[1] The postsurgical seizure outcome annually using the International League Against Epilepsy (ILAE) outcome scale[5] and noted the participants' medication. To improve accuracy, data on the timing of ASM withdrawal was corroborated by retrospectively reviewing medical notes and extracting the exact timing of the beginning of withdrawal and seizure relapses.

The Cleveland, Ohio, United States, retrospective cohort included individuals who underwent resective epilepsy surgery from January 2006 through June 2011 and met eligibility requirements (e.g., initiated ASM withdrawal, no post-operative seizure prior to ASM withdrawal, and at least a 1-year follow-up; n=117). Clinical data and outcomes were captured via medical chart review by epilepsy fellows and the clinical data manager. The primary outcome was time to seizure recurrence other than auras (i.e., ILAE outcome Class 3 or worse) after the beginning of anti-seizure medication withdrawal

For the Oxford, United Kingdom, cohort, individuals were selected from the Oxford University Hospital Epilepsy Surgery Registry from 2010 – 2020 (n=82).  Clinical data was extracted from the electronic case notes from individuals who met the study eligibility criteria (n=42).  Follow up was performed from a combination of face-to-face and telephonic consultations.  These follow-ups were performed mainly by the neurologist responsible for the patient's care.  In patients who were discharged from or left the Oxford Epilepsy Surgery service, follow-up information was provided by the General Practitioner who is responsible for their care.

The Lisbon, Portugal, cohort was selected from a cohort of 164 individuals that were included in the epilepsy surgery program of Hospital de Santa Maria between 1998 and 2009, who consented to participate and were 18 years or older at time of surgery[6]. Those that were not Engel class Ia for at least 12 months after surgery were excluded (n=97). All participants were postoperatively followed up by a neurologist with epilepsy expertise. Standardised phone interviews were used to assess for the frequency of seizures at the time of the interview, number of ASMs before surgery and at the time of the interview, ASM changes after surgery, timing for ASM changes after surgery and seizure recurrence.

The Bogota, Colombia, cohort included individuals from a prospective cohort of 232 adults with refractory epilepsy that had resective epilepsy surgery between 2013 and 2019. Those who had multiple interventions (n=26), who were not followed up for at least 1 year after surgery (n=103), who did not achieve freedom from non-aura seizures before ASM withdrawal (n=38) or did not start ASM withdrawal (=25) were excluded. All participants were followed up by epileptologists who interviewed them on postoperative seizures and ASM status. A data manager performed phone interviews.

In Cape Town, South Africa, individuals were selected from JTB Neurology Practice at the Constantiaberg Hospital (n=606). All patients who had received epilepsy surgery under JB care were reviewed and clinical data was extracted from the electronic case notes from patients who met the study eligibility criteria (n=105). Follow up was performed from a combination of face-to-face and telephonic consultations.  These follow-ups were performed solely by the neurologist responsible for the patient's care (JTB).

The Cardiff, United Kingdom, cohort was composed of consecutive patients selected from a clinical database (n=94), between March 1999 to December 2019. Baseline and follow up data were obtained from medical record. Those who had no medical records (n=6) or that did not fulfil inclusion criteria (n=44) were excluded.

The Melbourne, Australia, cohort included individuals who had epilepsy surgery at The Royal Melbourne Hospital (n=161, of who14 were excluded due to reintervention) or The Alfred Hospital (n=14) between 2000-2016. Medical records were reviewed and only those who fulfil the inclusion criteria were selected (n=35, n=9 respectively) for inclusion in the analysis.

The Shenzhen, China, cohort individuals were selected from the multicenter epilepsy surgery league program from April 2003 – May 2018 (n=350). Medical records were reviewed and only those who fulfil the inclusion criteria were selected (n=83) for the analysis. Follow up was performed from a combination of face-to-face and telephone or video consultations.

#### Supplementary Table 1. Characteristics of the cohorts

| Cohort | Overall (n=850) | London (n=350) | Cleveland (n=98) | Cape Town (n=105) | Shenzhen (n=83) | Melbourne (n=48) | Oxford (n=40) | Cardiff (n=44) | Lisbon (n=42) | Bogota (n=40) |
|---|---|---|---|---|---|---|---|---|---|---|
| **Demographics and past medical history** | | | | | | | | | | |
| Female sex | 447 (53) | 186 (53) | 50 (51) | 65 (62) | 29 (35) | 23 (48) | 21 (48) | 30 (68) | 22 (52) | 20 (50) |
| History of febrile seizures | 302 (37) | 186 (53) | 19 (26) | 18 (17) | 17 (20) | 6 (13) | 26 (65) | 19 (44) | 6 (17) | 5 (12) |
| Learning disability | 71 (9) | 17 (4.9) | 6 (8) | 6 (5) | 31 (37) | 2 (4) | 0(0) | 0 (0) | 5 (14) | 4 (10) |
| Psychiatric comorbidity | 225 (27) | 152 (43) | 12 (17) | 16 (15) | 6 (7) | 6 (12) | 15 (38) | 8 (18) | 10 (28) | 0 (0) |
| Age at epilepsy onset *(years)* | 13 [5-21] | 11 [4-18] | 10 [3-24] | 18 [8-28] | 15 [7-20] | 18 [9 -26] | 24 [16-32] | 12 [7-23] | 9 [4-18] | 13 [4-19] |
| Age at surgery *(years)* | 33 [25-44] | 33 [27-43] | 35 [20-49] | 37 [29-45] | 25 [22-31] | 39 [30-50] | 33 [28-44] | 34 [28-48] | 38 [32-45] | 30 [23-40] |
| History of focal to bilateral tonic clonic seizures before surgery | 586 (69) | 236 (67) | 74 (76) | 73 (69) | 57 (68) | 33 (70) | 22 (55) | 35 (79) | 19 (45) | 37 (92) |
| Frequency of seizures before surgery *(ordinal scale)* | 2 [2-3] | 2 [1-2] | 3 [2-3] | 3 [2-3] | 2 [2-2] | 2 [2-3] | 3 [2-4] | 3 [3-4] | 4 [2-4] | 4 [3-4] |
| Nonlesional MRI before surgery | 81 (10) | 15 (4) | 13 (13) | 35 (33) | 11 (13) | 2 (4) | 2 (5) | 0 (0) | 0 (0) | 3 (7) |
| Number of ASMs at surgery | 2 [2-3] | 2 [2-3] | 2 [2-2] | 2 [2-3] | 2 [1-2] | 2 [2-3] | 3 [2-3] | 2 [2-3] | 2 [2-3] | 3 [2-3] |
| Duration epilepsy at surgery *(years)* | 19 [9-29] | 21 [13-29] | 12 [4-31] | 17 [10-28] | 11 [6-20] | 21 [9-31] | 8 [5-14] | 22 [15-32] | 30 [14-39] | 18 [11-29] |
| **Surgical** | | | | | | | | | | |
| Laterality | | | | | | | | | | |
| Right | 394 (46) | 163 (46) | 52 (43) | 53 (51) | 31 (37) | 29 (60) | 20 (50) | 11 (25) | 23 (55) | 22 (56) |
| Left | 442 (53) | 187 (53) | 56 (57) | 51 (49) | 44 (53) | 19 (39) | 20 (50) | 33 (75) | 19 (45) | 17 (43) |
| Bilateral | 8 (1) | 0 (0) | 0 (0) | 0 (0) | 8 (9) | 0 (0) | 0 (0) | 0 (0) | 0 (0) | 0 (0) |
| Location | | | | | | | | | | |
| Temporal | 749(78) | 313 (89) | 98 (100) | 89 (85) | 64 (77) | 40 (83) | 38 (95) | 36 (82) | 41 (98) | 30 (75 |
| Extratemporal | 101 (22) | 37 (11) | 0 (0) | 16 (15) | 19 (23) | 8 (16) | 2 (5) | 8 (18) | 1 (2) | 10 (25) |
| Extent | | | | | | | | | | |
| Lesionectomy | 163 (22) | 41 (11) | 1 (1) | 10 (9) | 23 (27) | 14 (29) | 5 (13) | 11 (25) | 42 (100) | 17 (43) |
| Lobectomy | 575 (76) | 302 (86) | 97 (99) | 95 (90) | 54 (65) | 34 (71) | 35 (85) | 33 (75) | 0 (0) | 22 (56) |
| Hemispherectomy | 13 (1.8) | 7 (2) | 0 (0) | 0 (0) | 6 (7) | 0 (0) | 0 (0) | 0 (0) | 0 (0) | 0 (0) |
| Incomplete lesion removal | 92 (12) | 0 (0) | 34 (34) | 0 (0) | 21 (25) | 10 (24) | 3 (8) | 2 (4) | 22 (52) | 0 (0) |
| Pathology | | | | | | | | | | |
| Hippocampal sclerosis | 488 (58) | 247 (70) | 16 (16) | 41 (39) | 40 (48) | 25 (52) | 27 (71) | 31 (70) | 37 (88) | 24 (65) |
| Focal cortical dysplasia | 88 (10) | 11 (3) | 10 (10) | 9 (8) | 22 (26) | 14 (29) | 2 (5) | 3 (7) | 9 (21) | 8 (21) |

| | | | | | | | | | | |
|---|---|---|---|---|---|---|---|---|---|---|
| Dysembryoblastic neuroepithelial tumour | 51 (6) | 39 (11) | 1 (0) | 1 (1) | 0 (0) | 3 (6) | 6 (15) | 0 (0) | 1 (2) | 0 (0) |
| Cavernoma | 56 (8) | 27 (7) | 0 (0) | 5 (4.8) | 8 (9) | 6 (12) | 4 (10) | 4 (9) | 0 (0) | 2 (5) |
| Glioma | 31 (4) | 10 (3) | 6 (6) | 1 (1) | 2 (2) | 3 (6) | 1 (3) | 2 (4) | 2 (5) | 4 (11) |
| Dual pathology | 48 (6) | 10 (3) | 1 (0) | 2 (2) | 14 (17) | 6 (12) | 4 (10) | 0 (0) | 11 (26) | 0 (0) |
| Other | 126 (15) | 23 (6) | 28 (29) | 39 (37) | 23 (27) | 2 (4) | 2 (5) | 4 (9) | 5 (12) | 0 (0) |
| Normal | 22 (2) | 0 (0) | 1 (0) | 15 (14) | 3 (3) | 0 (0) | 0 (0) | 1 (2) | 0 (0) | 2 (5) |
| **Postsurgical** | | | | | | | | | | |
| Acute postsurgical seizures | 73 (10) | 24 (7) | 19 (19) | 2 (1.9) | 10 (12) | 2 (4) | 13 (33) | 3 (7) | 0 (0) | 0 (0) |
| Auras before ASM withdrawal | 87 (10) | 22 (6) | 19 (19) | 9 (8) | 8 (9) | 0 (0) | 7 (18) | 6 (13) | 8 (19) | 8 (20) |
| Non-aura seizure following ASM withdrawal | 270 (32) | 100 (28) | 64 (41) | 38 (36) | 18 (21) | 11 (23) | 5 (13) | 23 (52) | 17 (40) | 13 (33) |
| Time between surgery and beginning of ASM withdrawal *(years)* | 1 [0-2] | 1.5 [0.92-3] | 0.5 [0-2] | 0.5 [0-1] | 1 [1-2] | 0.95 [0-2] | 5.4 [4-7] | 1.7 [1-2] | 2 [2-4] | 0.7 [0-2] |
| Follow up duration *(years)* | 6 [3-10] | 10 [6-16] | 3 [3-4] | 4 [2-7] | 7 [5-9] | 5 [3-7] | 6 [5-7] | 5[4-7] | 6 [4-7] | 2 [1-3] |

Data displayed as N (%) or median [interquartile range]. ASM, antiseizure medication; MRI, magnetic resonance imaging; Preoperative seizure frequency categorized on an ordinal scale as 0 = no seizures or less than once a year, 1 = at least once a year, 2 = at least once a month, 3 = at least once a week, 4 = daily seizures. Incomplete lesion removal was determined when the epileptogenic zone included eloquent areas that were not subject to surgical resection

#### Supplementary Table 2. Univariable Cox regression analysis (primary model)

Univariable Cox regression analysis of predictors associated with time to non-aura seizure recurrence after beginning of ASM withdrawal following epilepsy surgery in the derivation cohort (London, UK)

| | **HR (95% CI)** | **P value** |
|---|---|---|
| **Demography** | | |
| Sex | | |
| Male | Reference | |
| Female | 1.09 (0.67-1.78) | 0.72 |
| Age at onset of epilepsy *(per 10 years)* | 0.92 (0.71-1.20) | 0.54 |
| Duration of epilepsy prior to surgery *(per 10 years)* | 1.18 (0.98-1.44) | 0.08 |
| Age at surgery *(per 10 years)* | 1.20 (0.97-1.50) | 0.14 |
| **Presurgical clinical history** | | |
| History of febrile seizures | 0.73 (0.45-1.85) | 0.43 |
| History of focal to bilateral tonic clonic seizures | 1.54 (0.87-2.70) | 0.13 |
| Presurgical seizure frequency *(per month)* | 0.99 (0.99-1.00) | 0.82 |
| History of learning disability | 0.84 (0.20-3.45) | 0.81 |
| History of any psychiatric comorbidity | 0.91 (0.55-1.50) | 0.72 |
| Number of ASMs at time of surgery | 1.24 (0.95-1.60) | 0.10 |
| Normal pre-surgical MRI | 0.86 (0.21-3.52) | 0.80 |
| **Surgery** | | |
| Side of resection | | |
| Left | Reference | |
| Right | 0.90 (0.55-1.47) | 0.70 |
| Type of resection | | |
| Lobectomy | Reference | |
| Lesionectomy | 0.78 (0.33-1.80) | 0.57 |
| Hemispherectomy | 0.64 (0.08-4.66) | 0.66 |
| Location of resection | | |
| Temporal | Reference | |
| Extratemporal | 1.11 (0.55-2.51) | 0.76 |
| Incomplete resection | <0.01 (0-Inf) | 0.99 |
| Pathology | | |
| Hippocampal sclerosis | 1.18 (0.69-2.04) | 0.52 |
| Focal cortical dysplasia | 0.52 (0.07-3.79) | 0.52 |
| Dysembryoplastic neuroepithelial tumour | 0.82 (0.35-1.89) | 0.64 |
| Arteriovenous malformation | 0.96 (0.41-2.22) | 0.92 |
| Glioma | <0.01 (0-Inf) | 0.99 |
| Other | 1.22 (0.49-3.04) | 0.66 |
| **Postsurgical clinical history** | | |
| Acute postsurgical seizures | 1.34 (0.58-3.10) | 0.49 |
| Auras after surgery before beginning of ASM withdrawal | 3.6 (1.81-7.05) | **<0.0001** |
| Time to begin ASM withdrawal *(years from surgery)* | 0.93 (0.85-1.01) | 0.10 |

Supplementary Table 3. Internal-external cross-validation (primary model)

| Omitted cohort | Model discrimination in remaining studies |
|---|---|
| London | 0.67 |
| Cleveland | 0.65 |
| Cape Town | 0.65 |
| Shenzhen | 0.66 |
| Oxford | 0.65 |
| Melbourne | 0.66 |
| Lisbon | 0.65 |
| Colombia | 0.65 |

For each run one cohort was omitted after which the model was fitted on the remaining studies and a c-statistic was computed. C-statistic was corrected by bootstrapping. Empty cells are due to singularity of the data.

#### Supplementary Table 4. Univariable Cox regression analysis (secondary model)

Univariable Cox regression analysis of predictors associated with time to any seizure recurrence after beginning of ASM withdrawal following epilepsy surgery in the complete seizure free cohort*

| | **HR (95% CI)** | **P value** |
|---|---|---|
| **Demography** | | |
| Sex | | |
| Male | Reference | |
| Female | 1.22 (0.94-1.59) | 0.13 |
| Age at onset of epilepsy *(per 10 years)* | 1.02 (0.91-1.14) | 0.66 |
| Duration of epilepsy prior to surgery *(per 10 years)* | 1.13 (1.04-1.24) | 0.015 |
| Age at surgery *(per 10 years)* | 1.17 (1.05-1.31) | 0.003 |
| **Presurgical clinical history** | | |
| History of febrile seizures | 0.75 (0.57-0.99) | 0.044 |
| History of focal to bilateral tonic clonic seizures | 1.80 (1.31-2.45) | <0.001 |
| Presurgical seizure frequency *(per month)* | 1.32 (1.1§4-1.52) | <0.001 |
| History of any psychiatric comorbidity | 0.72 (0.53-0.97) | 0.046 |
| Number of ASMs at time of surgery | 1.16 (0.95 -1.29) | 0.154 |
| Normal pre-surgical MRI | 1.57 (1.03-2.40) | 0.032 |
| **Surgery** | | |
| Side of resection | | |
| Left | Reference | |
| Right | 1.00 (0.78-1.28) | 0.979 |
| Type of resection | | |
| Lobectomy | Reference | |
| Lesionectomy | 0.85 (0.58-1.23) | 0.394 |
| Hemispherectomy | 0.35 (0.09-1.41) | 0.141 |
| Location of resection | | |
| Temporal | Reference | |
| Extratemporal | 0.76 (0.50-1.16) | 0.218 |
| Incomplete resection | 1.95 (1.26-3.01) | 0.002 |
| Pathology | | |
| Hippocampal sclerosis | 0.99 (0.76-1.30) | 0.983 |
| Focal cortical dysplasia | 0.93 (0.57-1.54) | 0.803 |
| Dysembryoplastic neuroepithelial tumour | 0.75 (0.44-1.27) | 0.28 |
| Arteriovenous malformation | 0.71 (0.42-1.20) | 0.203 |
| Glioma | 1.05 (0.46-2.38) | 0.89 |
| Other | 1.01 (0.68-1.49) | 0.961 |
| **Postsurgical clinical history** | | |
| Acute postsurgical seizures | 0.71 (0.40-1.28) | 0.264 |
| Time to begin ASM withdrawal *(years from surgery)* | 0.86 (0.80-0.92) | <0.001 |

*no auras between surgery and ASM withdrawal.

#### Supplementary Table 5. Multivariable Cox regression analysis (secondary model)

Multivariable Cox regression analysis of time to any seizure recurrence after beginning of ASM withdrawal following epilepsy surgery.

| **Predictors** | **aHR (95% CI)** | **p-value** | **Δ AIC*** |
|---|---|---|---|
| Time to beginning of ASM withdrawal *(per year from surgery)* | 0.85 (0.79-0.92) | <0.001 | -7.1 |
| Focal to bilateral tonic- clonic seizures before surgery | 1.88 (1.37-2.59) | <0.001 | -4.6 |
| Presurgical seizure frequency *(as an ordinal scale)* | 1.29 (1.12-1.50) | <0.001 | -6.2 |
| Duration of epilepsy before surgery (per 10 years) | 1.13 (1.02-1.25) | 0.011 | -1.6 |
| History of febrile seizures | 0.80 (0.60-1.05) | 0.116 | -0.52 |
| Normal pre-surgical MRI | eliminated step 4 | 0.59 | 0.22 |
| Age at surgery (per ten years) | eliminated step 3 | 0.60 | 0.77 |
| Hippocampal sclerosis on neuropathology | eliminated step 2 | 0.67 | 0.83 |
| Number of ASMs at time of surgery | eliminated step 1 | 0.50 | 1.55 |

N=639. aHR, adjusted hazard ratio; MRI, magnetic resonance imaging; ΔAIC=change in Akaike information criterion after elimination of a variable at each step; * A negative value implies that the variable improves the model and should be kept in the model.

#### Supplementary Table 6. Univariable Cox regression analysis (tertiary model)

Univariable Cox regression analysis of predictors associated with time to any seizure recurrence after beginning of ASM withdrawal following epilepsy surgery including only those who had temporal lobe surgery and were completely seizure free before withdrawal*.

| | HR (95% CI) | P value |
|---|---|---|
| **Demography** | | |
| Sex | | |
| Male | Reference | |
| Female | 1.21 (0.94-1.58) | 0.134 |
| Age at onset of epilepsy *(per 10 years)* | 1.06 (0.93-1.15) | 0.471 |
| Duration of epilepsy prior to surgery *(per 10 years)* | 1.10(0.98-1.18) | 0.096 |
| Age at surgery *(per 10 years)* | 1.15(1.08-1.27) | 0.010 |
| **Presurgical clinical history** | | |
| History of febrile seizures | 0.70 (0.52-0.94) | 0.015 |
| History of focal to bilateral tonic clonic seizures | 2.00 (1.47-1.58) | <0.001 |
| Presurgical seizure frequency *(per month)* | 0.99 (0.95-2.73) | <0.001 |
| History of any psychiatric comorbidity | 0.71 (0.52-0.96) | 0.039 |
| Number of ASMs at time of surgery | 1.48 (0.90 -2.43) | 0.116 |
| Normal pre-surgical MRI | 1.47 (0.97-2.12) | 0.064 |
| **Surgery** | | |
| Side of resection | | |
| Left | Reference | |
| Right | 1.03 (0.83-1.36) | 0.531 |
| Type of resection | | |
| Lobectomy | Reference | |
| Lesionectomy | 1.17 (0.78-1.77) | 0.429 |
| Hemispherectomy | 0.55 (0.07-3.62) | 0.559 |
| Incomplete resection | 1.43 (0.96-2.13) | 0.041 |
| Pathology | | |
| Hippocampal sclerosis | 0.93 (0.73-1.27) | 0.612 |
| Focal cortical dysplasia | 0.74 (0.43-1.21) | 0.270 |
| Dysembryoplastic neuroepithelial tumour | 0.68 (0.31-1.19) | 0.183 |
| Arteriovenous malformation | 1.02 (0.67 -1.56) | 0.910 |
| Glioma | 0.98 (0.78-2.40) | 0.755 |
| Other | 1.12 (0.68-1.61) | 0.520 |
| **Postsurgical clinical history** | | |
| Acute postsurgical seizures | 0.58 (0.33-1.05) | 0.080 |
| Time to begin ASM withdrawal *(years from surgery)* | 0.88 (0.82-0.94) | <0.001 |

*no auras between surgery and ASM withdrawal.

#### Supplementary Table 7. Multivariable Cox regression analysis (tertiary model)

Multivariable Cox regression analysis of time to any seizure recurrence after beginning of ASM withdrawal following epilepsy surgery including only those who had temporal lobe surgery and were completely seizure free before withdrawal.

| **Predictors** | **aHR (95% CI)** | **p-value** | **Δ AIC*** |
|---|---|---|---|
| Time to beginning of ASM withdrawal *(per year from surgery)* | 0.85 (0.79-0.92) | <0.001 | -19.68 |
| Focal to bilateral tonic- clonic seizures before surgery | 1.94 (1.38-2.74) | 0.0001 | -15.28 |
| Incomplete surgery | 1.86 (1.17-2.96) | 0.008 | -2.08 |
| Duration of epilepsy before surgery (per 10 years) | 1.10 (0.99-1.22) | 0.069 | -1.58 |
| Psychiatric comorbidity | 0.73 (0.53-1.00) | 0.054 | -0.18 |
| History of febrile seizures | eliminated step 4 | 0.13 | 0.02 |
| Normal pre-surgical MRI | eliminated step 3 | 0.22 | 0.62 |
| Acute postsurgical seizures | eliminated step 2 | 0.50 | 1.55 |
| Age at surgery (per ten years) | eliminated step 1 | 0.52 | 1.64 |

The model showed an optimism-corrected *c* statistic of 0.68 (95% CI 0.63-0.73)

N=558. aHR, adjusted hazard ratio; MRI, magnetic resonance imaging; ΔAIC=change in Akaike information criterion after elimination of a variable at each step; * A negative value implies that the variable improves the model and should be kept in the model.

#### Supplementary Table 8. Univariable Cox regression analysis (complete ASM withdrawal

Univariable Cox regression analysis of predictors associated with complete ASM withdrawal following epilepsy surgery including only those who were completely seizure free after surgery and follow up.

| | **HR (95% CI)** | **P value** |
|---|---|---|
| **Demography** | | |
| Sex | | |
| Male | Reference | |
| Female | 1.02 (0.76-1.33) | 0.844 |
| Age at onset of epilepsy *(per 10 years)* | 0.98 (0.81-1.05) | 0.224 |
| Duration of epilepsy prior to surgery *(per 10 years)* | 0.85 (0.75-0.96) | 0.010 |
| Age at surgery *(per 10 years)* | 0.74 (0.64-0.85) | <0.001 |
| **Presurgical clinical history** | | |
| History of febrile seizures | 0.80 (0.59-1.07) | 0.142 |
| History of focal to bilateral tonic clonic seizures | 0.77 (0.57-1.09) | 0.087 |
| Presurgical seizure frequency *(per month)* | 0.79 (0.66-0.94) | 0.010 |
| History of learning disability | 2.06(1.32-3.22) | 0.001 |
| History of any psychiatric comorbidity | 0.80 (0.62-1.03) | 0.181 |
| Number of ASMs at time of surgery | 0.68 (0.56-0.82) | 0.001 |
| Normal pre-surgical MRI | 0.72 (0.37-1.41) | 0.347 |
| **Surgery** | | |
| Side of resection | | |
| Left | Reference | |
| Right | 0.94 (1.06-1.24) | 0.67 |
| Extent | | |
| Temporal | Reference | |
| Extratemporal | 1.4 (0.92-2.12 | 0.113 |
| Type of resection | 1.25 (0.64-2.44) | 0.506 |
| Lobectomy | Reference | |
| Lesionectomy | 0.62 (0.43-0.88) | 0.009 |
| Hemispherectomy | 1.10 (0.51-2.36) | 0.801 |
| Incomplete resection | 1.23 (0.70-2.17) | 0.46 |
| Pathology | | |
| Hippocampal sclerosis | 0.79 (0.59-1.05) | 0.114 |
| Focal cortical dysplasia | 1.42 (0.88-2.30 | 0.145 |
| Dysembryoplastic neuroepithelial tumour | 1.24 (0.76-2.01) | 0.385 |
| Arteriovenous malformation | 1.40 (0.90-2.17) | 0.126 |
| Glioma | 4.08 (2.05-8.08) | <0.001 |
| Other | 0.81 (0.51-1.29) | 0.389 |
| **Postsurgical clinical history** | | |
| Acute postsurgical seizures | 0.84 (0.47-1.48) | 0.552 |
| Time to begin ASM withdrawal *(years from surgery)* | 0.80 (0.7150.87) | <0.001 |

N = 436.

#### Supplementary Table 9. Multivariable Cox regression analysis (complete ASM withdrawal)

Multivariable Cox regression analysis of predictors associated with complete ASM withdrawal following epilepsy surgery including only those who were completely seizure free after surgery and follow up.

| **Predictors** | **aHR (95% CI)** | **p-value** | **Δ AIC*** |
|---|---|---|---|
| Time to beginning of ASM withdrawal *(per year from surgery)* | 0.79 (0.73-0.86) | <0.001 | -47.8 |
| Age at time of surgery (per 10 years) | 0.76 (0.65-0.84) | <0.001 | -12.8 |
| Number of ASMs at time of surgery | 0.70 (0.58-0.86) | <0.001 | -10 |
| Learning disability | 0.67 (1.24-3.06) | 0.003 | -5.7 |
| Presurgical seizure frequency *(per month)* | 0.82 (0.68-0.98) | 0.037 | -4.5 |
| Glioma on pathology | 2.70 (1.28-5.71) | 0.008 | -4.00 |
| Duration of epilepsy before surgery (per 10 years) | eliminated step 2 | 0.080 | 0.46 |
| Surgery extent (Lesionectomy) | eliminated step 1 | 0.48 | 1.46 |

The model showed an optimism-corrected *c* statistic of 0.73 (95% CI 0.68-0.78)

aHR, adjusted hazard ratio; MRI, magnetic resonance imaging; ΔAIC=change in Akaike information criterion after elimination of a variable at each step. * A negative value implies that the variable improves the model and should be kept in the model

#### Supplementary Figure 1. Flowchart of participants included in the study

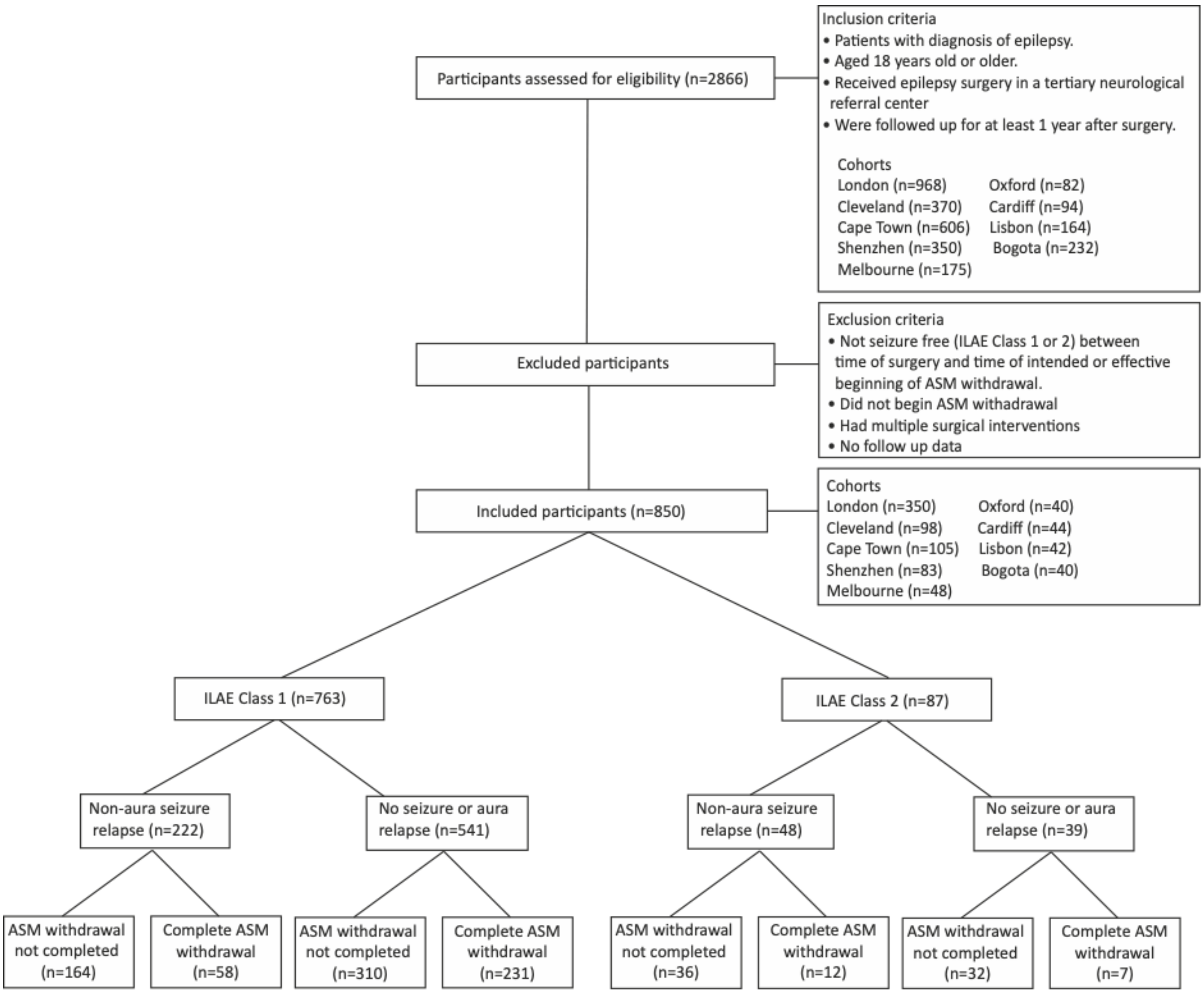

#### Supplementary Figure 2. Calibration plots

Calibration plots for the primary model, London cohort (A), all other cohorts (B), secondary model (C), tertiary model (D) and final model (E) at 2-years (left) and 4-years (right) after beginning of ASM withdrawal following epilepsy surgery. Vertical lines are 95% confidence intervals.

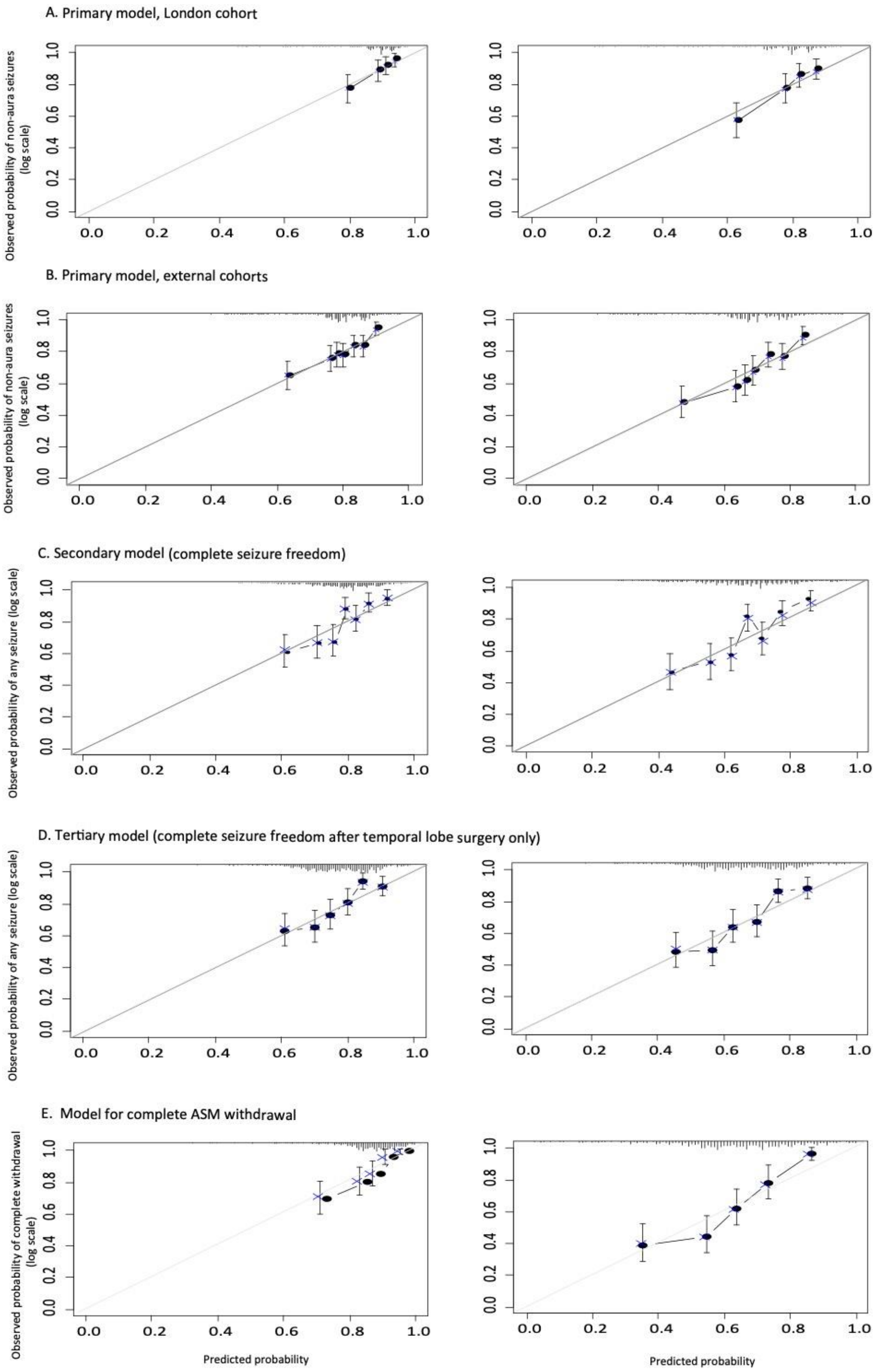

Supplementary Figure 3. Nomogram for predicting 2- and 4-year seizure outcome after beginning ASM withdrawal after temporal lobe surgery only.

Freedom from any seizure including auras after beginning ASM withdrawal after temporal lobe surgery. Instructions: Determine the individual risk in three steps: 1) For every variable on the left, count the points given at the top, 2) Add up the points to a total, 3) Determine the associated recurrence risk at 2 and 4 years according to the calculated point total.

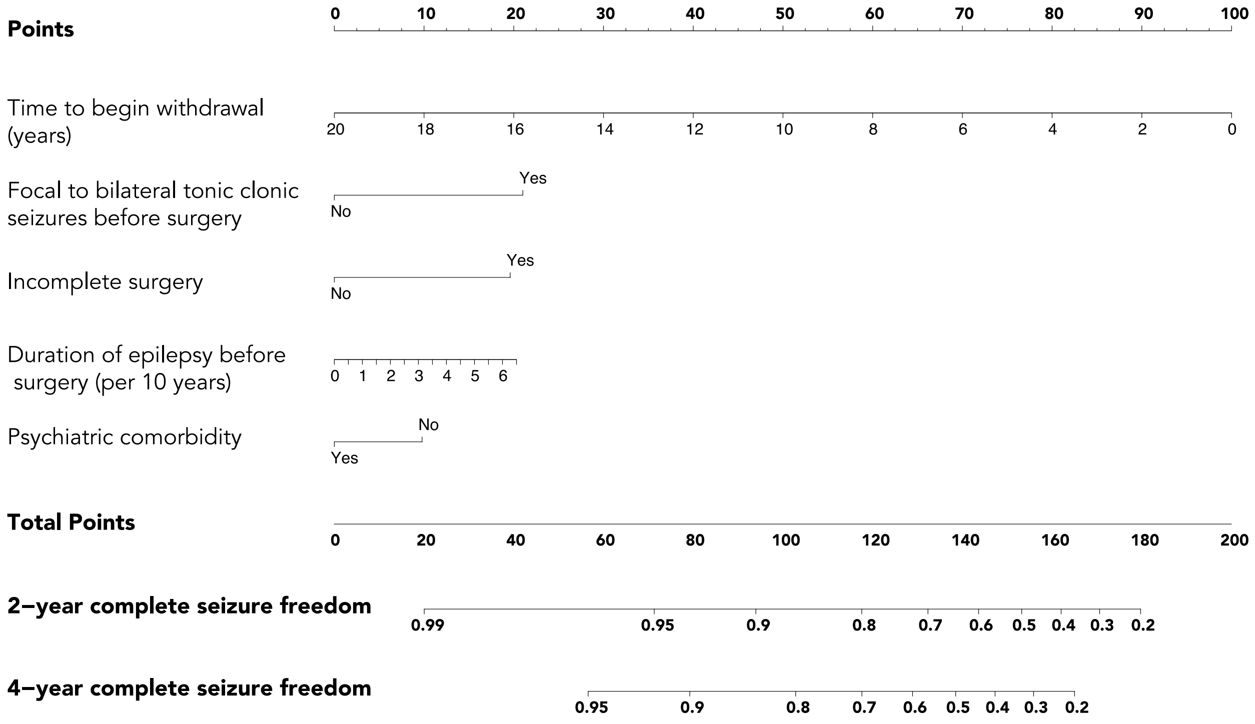

#### Supplementary Figure 4. Nomogram for predicting complete ASM withdrawal at 2- and 4-years after beginning ASM withdrawal.

Completing ASM withdrawal after epilepsy surgery for those who started withdrawal and have been seizure free since surgery.  Instructions: Determine the individual risk in three steps: 1) For every variable on the left, count the points given at the top, 2) Add up the points to a total, 3) Determine the associated risk at 2 and 4 years according to the calculated point total.

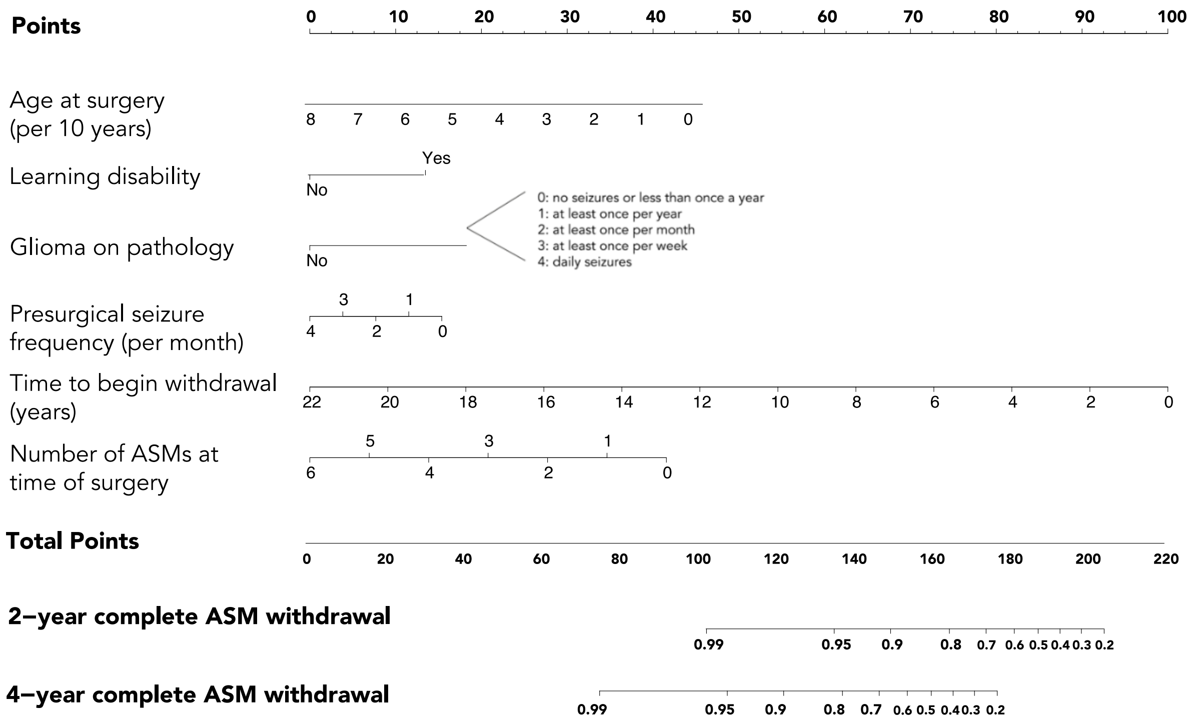

#### Supplementary Figure 5. Receiver Operator Characteristic curves (ROC)

ROC curves for the (A) primary model (non-aura seizure recurrence), (B) secondary model (any seizure recurrence), (C) tertiary model (any seizure recurrence, temporal lobe surgery) and (D) complete ASM withdrawal at (1) 2- and (2) 4-years after beginning ASM withdrawal.

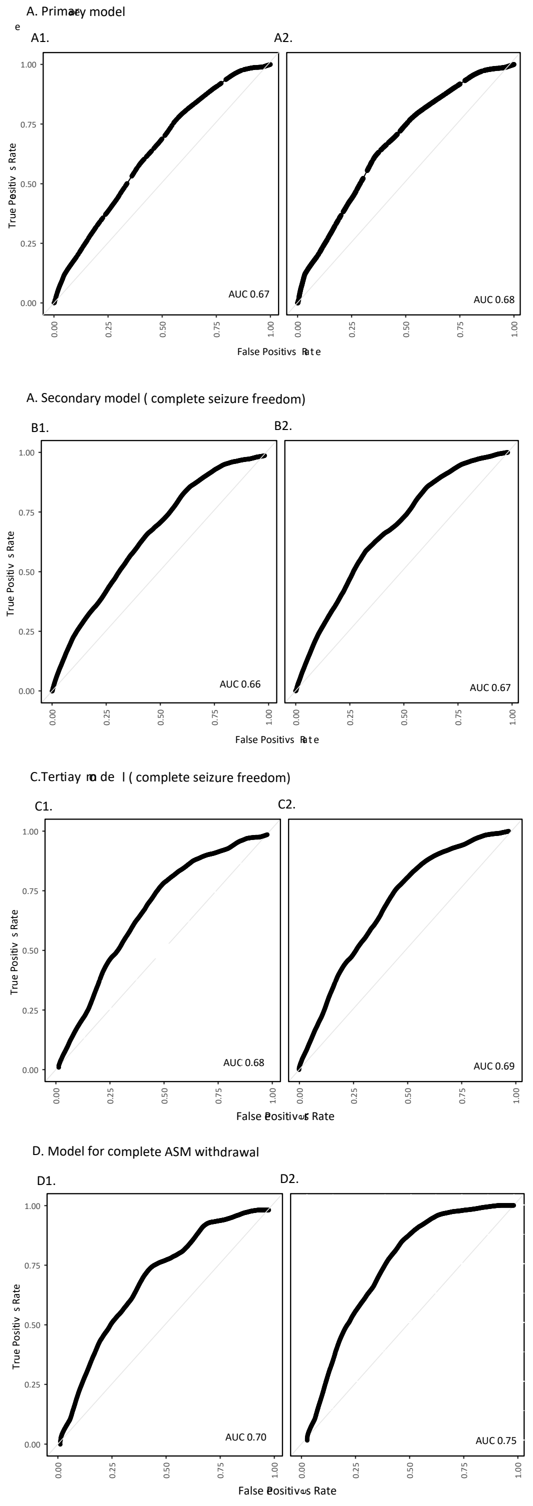
